# Optimizing vaccination strategies for mpox control in endemic areas: Modeling insights from the Democratic Republic of Congo

**DOI:** 10.1101/2025.08.07.25333225

**Authors:** Aakash Pandey, Ian Spicknall, Andrea M. McCollum, Christine M. Hughes, Beatrice Nguete, Toutou Likafi, Robert Shongo Lushima, Placide Mbala-Kingebeni, Joelle Kabamba, Didine Kaba, Yoshinori Nakazawa

## Abstract

The global mpox outbreak of 2022, caused by the Clade IIb strain of monkeypox virus, underscored the potential of this virus to pose a significant public health threat on a global scale. The Democratic Republic of Congo is currently facing multiple outbreaks associated with Clade I. Effectively controlling localized community transmission within endemic areas through vaccination can reduce the likelihood of broader regional or even global outbreaks. Large-scale community vaccination in DRC is challenged by limited resources, including vaccine availability during early outbreaks in remote areas, whereas limited surveillance, contact tracing, and accessibility to remote locations can reduce the effectiveness of targeted ring vaccination. Furthermore, recent outbreaks in DRC have been driven by both sexual and non-sexual close contact transmissions. Here, we used an agent-based model with stochastic transmission within and between households to assess the effectiveness of ring vaccination for controlling localized community transmission in the presence of incomplete case reporting and delay in vaccination. We consider both nonsexual close contact and sexual transmission. We found that ring vaccination, even with 25-50% reporting, is effective in reducing outbreak cluster sizes and the likelihood of large cluster sizes (>5 cases), particularly when implemented shortly after detection of initial cases. The effectiveness of ring vaccination reduces with the inclusion of sexual transmission. We show that outbreak size and the likelihood of large clusters are reduced when responding to every reported infection, even with 2-3 weeks of delay. Settings with strong surveillance systems characterized by high levels of reporting will have earlier case detection, enabling earlier response and improving the effectiveness of ring vaccination.

## Introduction

Mpox is an emerging zoonotic disease, caused by monkeypox virus (MPXV), that is endemic to Central and Western Africa. Clade I MPXV is found in central Africa, whereas Clade II MPXV is endemic to Western African countries; although geographically separated, both clades are present in Cameroon [1,2]. Historically, mpox outbreaks have led to small clusters with short chains of human-to-human transmission; however, it has been hypothesized that multiple lineages of MPXV clade II co-circulated undetected in humans for several years before it caused the global outbreak of 2022 [3]. Sexual or intimate physical contact was one of the primary routes by which MPXV clade IIb was transmitted during the global outbreak of 2022. In 2023-2024, the Democratic Republic of Congo reported significantly increased numbers of suspected cases associated with MPXV clade I and in areas of the country that do not normally have mpox. It then spread beyond the borders of the DRC into neighboring countries [4]. In eastern DRC, MPXV transmission is associated with sexual contacts in addition to close contact (i.e., non-sexual close physical contact) transmission as seen in previous MPXV clade I outbreaks [5–8]. Given that Clade I has historically been associated with a higher proportion of individuals with severe disease compared to Clade II [2,9], it is critical to understand how changing epidemiology impacts transmission control strategies.

Before 2022, the primary source of human MPXV infection has historically been from spillovers from wildlife, followed by limited transmission within households [10–13]; with the longest reported transmission chain between humans being seven generations [14]. Genomic data from samples collected between 2018-2024 from mpox-positive patients in 17 of the 26 provinces of DRC suggests multiple, independent zoonotic transmission events [15]. Responding to each spillover event with ring vaccination could effectively reduce the likelihood of larger outbreaks [16]; however, this requires an underlying surveillance system to detect these events early and response mechanisms to implement interventions quickly. This may not be viable in low- and middle-income countries (LMICs) where resources for surveillance and subsequent public health actions may be limited. The lack of a strong surveillance system, misdiagnosis of cases, and poor healthcare infrastructure can lead to underreporting [17–19]. A household study after a 2013 mpox outbreak in Bokungu health zone of Tshuapa province estimated a reporting rate of 41% [14]. This can reduce the effectiveness of ring vaccination. Additionally, even when a case is identified, finding close contacts of household members can be difficult due to several reasons, including stigma and vaccine hesitancy, among others [20,21]. Further, finding contacts and accessing households in remote locations with little to no healthcare infrastructure can delay ring vaccination implementation. Therefore, it is critical to understand the effect of these limitations on the efficacy of ring vaccination.

Here, we developed an agent-based model with individuals as ‘agents’ to understand the effectiveness of ring vaccination strategies when considering the effect of limited reporting, incomplete contact tracing, and delay in the start of ring vaccination implementation. We explored the effectiveness of four different ring vaccination scenarios. One scenario consists of ring vaccination within households with reported infections only; a second scenario considers additionally vaccinated non-household contacts. Similar scenarios are explored for populations that also have transmission through a commercial sex network. We explored how reporting thresholds for initiating a ring vaccination response affect ring vaccination effectiveness. Additionally, to inform decision making by public health officials, variable delays in response were incorporated, thereby identifying ring vaccination strategies that can be effective in settings with fewer resources and or weaker surveillance capacity.

## Methods

### Baseline disease model

We used a stochastic agent-based modeling to simulate mpox disease dynamics within and between households. Individual states included Susceptible-Exposed-Infected-Recovered-Dead (SEIRD) (Fig. 1). Each individual was assigned to a specific household. The state transition rates were determined by exponentially distributed probabilities. We considered two different models with different modes of transmission. In the first model, we assumed transmission through close physical contact without sexual contact – akin to the historical endemic transmission - where within-household contacts were assumed to be homogeneous, whereas between-household contacts were determined from an Erdős-Rényi random graph. In the second model, everything was identical to the first model except for the addition of sexual transmission through a sexual network composed of commercial sex workers and clients, akin to the transmission observed in Eastern DRC. In this model, 10% of households were assumed to include a commercial sex worker, whereas 40% of households were assumed to host at least one client. A dynamic network was created to simulate contacts between commercial sex workers and clients, assuming no turnover in commercial sex worker and client statuses. Details of the model and simulation process, including the addition of the ring vaccination intervention, are described below.

**Fig. 1.**
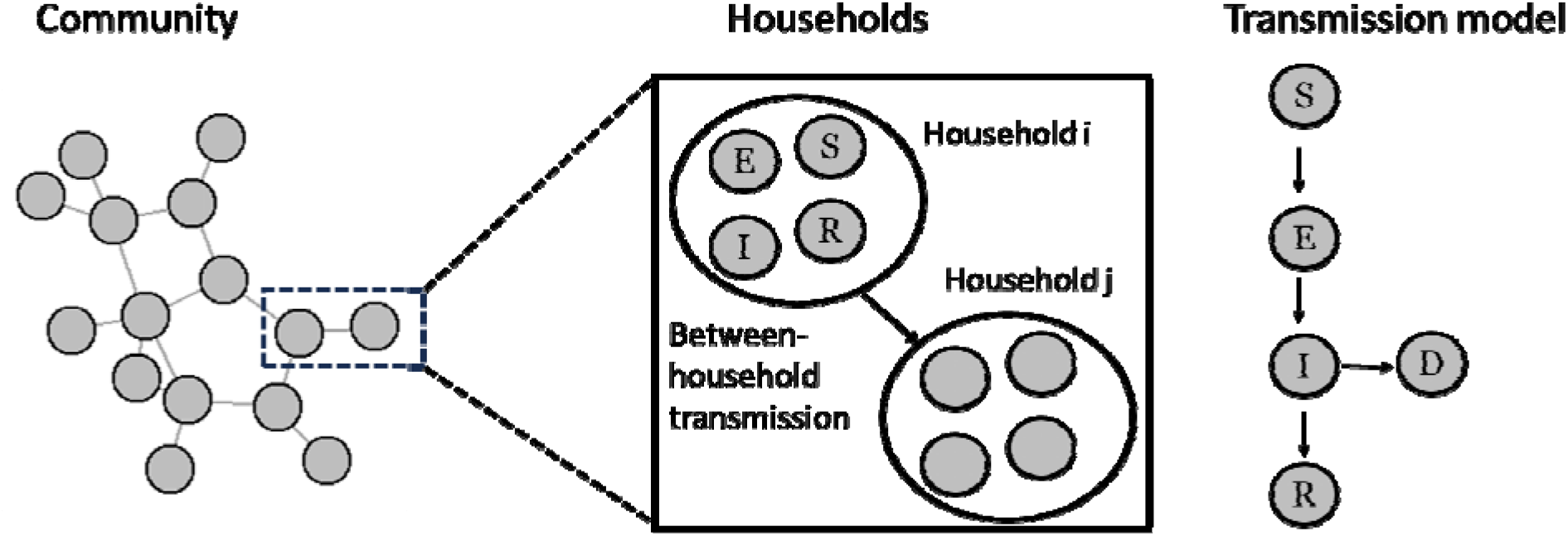
Mpox transmission model. A community network is made up of households that are composed of individuals. Between-household contacts are represented by edges connecting households. The transmission model shows the sequence of transitions for individuals. S, E, I, R, D represent susceptible, exposed, infected, recovered, and dead states of individuals, respectively.

### Disease transmission

In the first model without sexual transmission, we assumed all household members interact homogeneously and transmission within household members is driven by close contact among them. Contacts between members of different households were modeled through a static Erdős-Rényi random graph. The graph was generated using ‘igraph’ package in R software [22]. The probability of edge formation between two households was estimated by fitting the baseline transmission model to data as described in the following section. Both transmission rates within households and between households were estimated by fitting the model to the surveillance data from Tshuapa, DRC [10] and were assumed to be exponentially distributed.

In the second model that adds sexual transmission to the first model, we added a dynamic transmission network between commercial sex workers and clients [5]. The edges between commercial sex workers and clients were assigned randomly, and the number of edges per day followed a Poisson distribution with a mean of 2. Although the number of clients per day can vary substantially and little is known about such networks in the mining town of South Kivu, similar estimates were previously found in Kinshasa [23]. A new set of edges (i.e., sexual contacts) was generated for each time step (i.e., daily). We assumed the maximum number of sexual contacts per day was at 1 for clients; for a CSW, it was determined by random sampling from a Poisson distribution with a mean of 2. Since we do not have a good understanding of the transmissibility of clade I mpox through sexual contact, the per day per contact probability of transmission between CSW and client was calibrated to be 0.125 to ensure that an outbreak occurs in the sexual network.

### Model fitting

Disease parameters were estimated by fitting the baseline model with close contact transmission within and between households to the surveillance data from Tshuapa province collected during 2013-2017. Details of the data collection process and its analysis are already published [10]. Although more data is available, we focused on using the data from this period as it represents a relatively stable period of surveillance efforts. Parameters were fitted using the approximate Bayesian computation method [24]. Specifically, we simulated the model 1000 times with parameters selected from uniform distributions and the output of the cluster size distribution was fitted with the estimated frequency of cluster sizes obtained from the data. All the parameters and their fitted median values are listed in Table 1.

**Table 1.** Parameters and median values used in the model.

| Definition | Value | Source |
| --- | --- | --- |
| Transmission probability within a household | 0.013/day | Estimated (Fig. S1) |
| Transmission probability between households | 0.0048/day | Estimated (Fig. S1) |
| Incubation period | 7 days | Estimated (Fig. S1) |
| Infectious period | 23 days | Estimated (Fig. S1) |
| Probability of edge formation between households | 0.01 | Estimated |
| Probability of death due to infection | 0.004 | Beer and Rao, 2019[28] |
| Vaccine effectiveness | 0.66 | Deputy et al., 2023 [25] |
| Transmission probability per sexual contact | 0.125 | Assumed |

### Vaccination

Ring vaccination was applied based on the available information on within and between household contacts, as well as the reporting rate. Our assumed decision tree for triggering ring vaccination is shown in Fig. 2. When an infection occurs in a household, we assume that the infection is reported in the household with probability *r*_*p*_. We assumed reporting probabilities of either 25% or 50%, a range that captures the observed reporting rate from the Tshuapa province [10,14]. If an infection is not reported in the household, no response is triggered. If the total number of reported infections crosses a pre-defined threshold (either 1, 2 or 3 cases), ring vaccination is initiated. For example, when the threshold is 1, we initiate ring vaccination for every reported infection. If the threshold is 2, we only respond when there are 2 reported infections, and so on. Responding to reports of infections may not be immediate, depending on the health care infrastructure and accessibility of households; thus, we introduce different degrees of delays. We consider 1, 2 and 3 weeks of delays and compare them with the baseline of immediate response. Those who get infected during the delay period are not eligible for vaccination. Hence, only individuals who are in susceptible and exposed states after the delay period can be vaccinated. We assume a one-dose vaccine with an effectiveness of 67% [25] and assume a ‘leaky’ mechanism, where a vaccinated individual could be infected with repeated exposures, but does not contribute to further transmission. Although post-exposure vaccine effectiveness can vary with time since exposure [26], we assume a similar level of protection for both exposed and susceptible individuals.

**Fig. 2.**
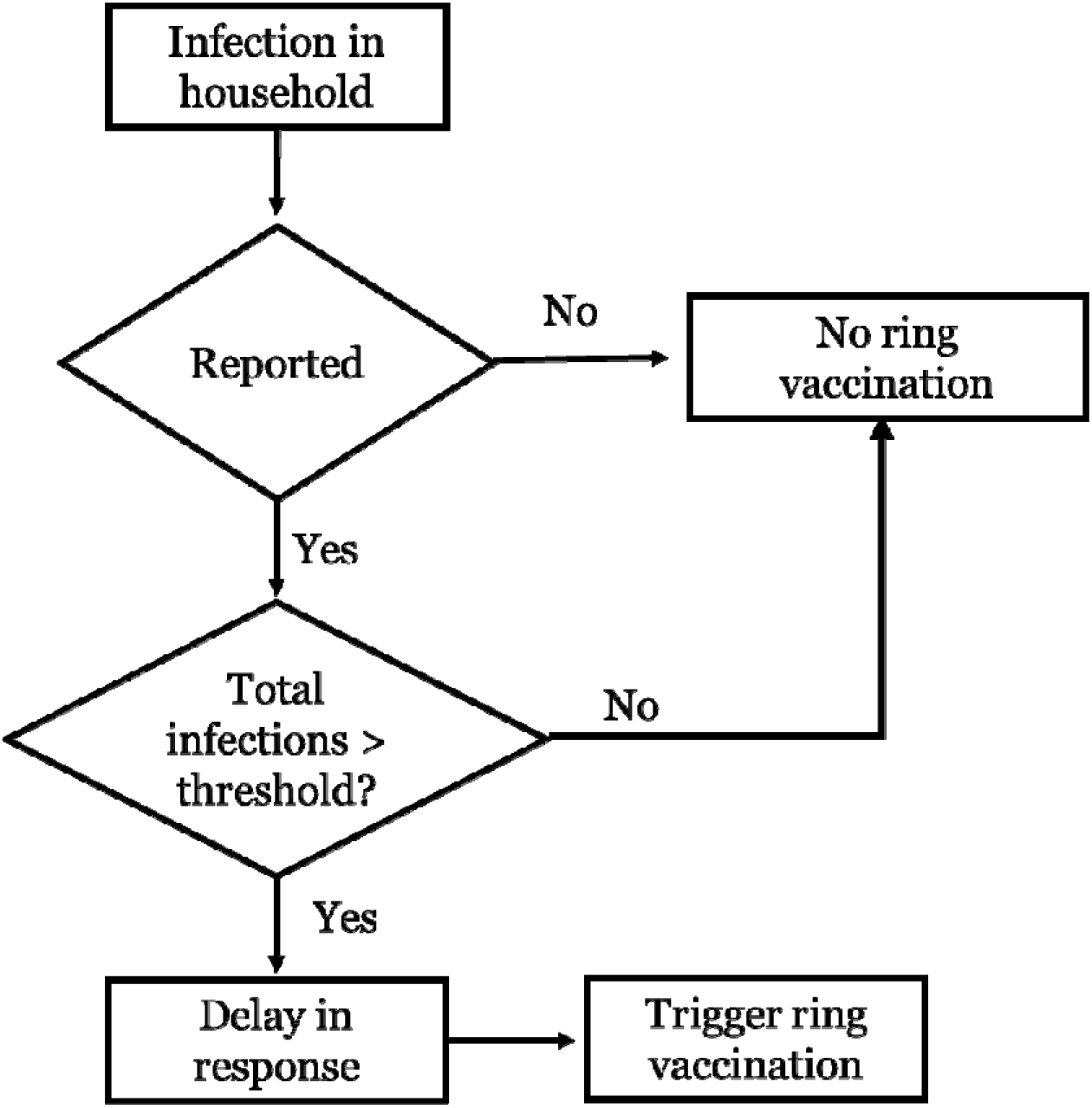
Decision tree for initiation of ring vaccination.

To account for different degrees of availability of contact information for ring vaccination, we consider 2 scenarios: one with vaccination only of households with reported infections (i.e., assuming between household contacts cannot be identified); and another including between household contacts in the ring vaccination (i.e., assuming between household contacts can be identified). We explored these two scenarios in both the non-sexual transmission model and the model that adds sexual transmission. We further assume that sexual contacts in commercial sexual networks cannot be identified.

### Simulation and measured outcomes

We started with one infected individual in a random household to mimic spillover or case importation events. For each scenario, we ran 1000 simulations and measured the probability of large outbreaks following ring vaccination. For the model with close contact transmission only, we summarized the probabilities of outbreak size being greater than 5 following introductions of a single case in a household. The average household size in the DRC is 5.3 [27]. The distribution of probabilities for each scenario was calculated assuming a binomial distribution based on the number of simulations that crossed the outbreak size of 5 infections. We also calculated relative risk reduction due to vaccination of additional between-household contacts using the following formula:

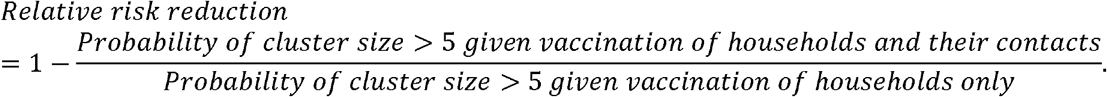

For the second model that adds sexual transmission, we randomly seeded an infection in a household with a commercial sex worker. Like the previous model, we measured the probability of large clusters and the relative risk reduction with additional vaccination between household contacts of infected households. Since transmission in a sexual network can result in a large cluster, we also measure the mean cluster size and the relative reduction in mean cluster size for two vaccination scenarios.

## Results

### Vaccinating only reported households in the non-sexual transmission model

With 25 % reporting, there was a small reduction in the probability of cluster size being greater than 5 when ring vaccination is initiated for every reported infection (i.e., threshold=1) (Fig. 3a). No difference was observed regardless of reporting thresholds or delay period length to initiate ring vaccination. With 50% reporting, there was a larger reduction in the probability of cluster size being greater than 5. For reporting thresholds of 1 and 2 cases, the probabilities of large cluster formation increased with delays in ring vaccination initiation. For a threshold of 3, we did not see any difference from the no-vaccination scenario.

**Fig. 3.**
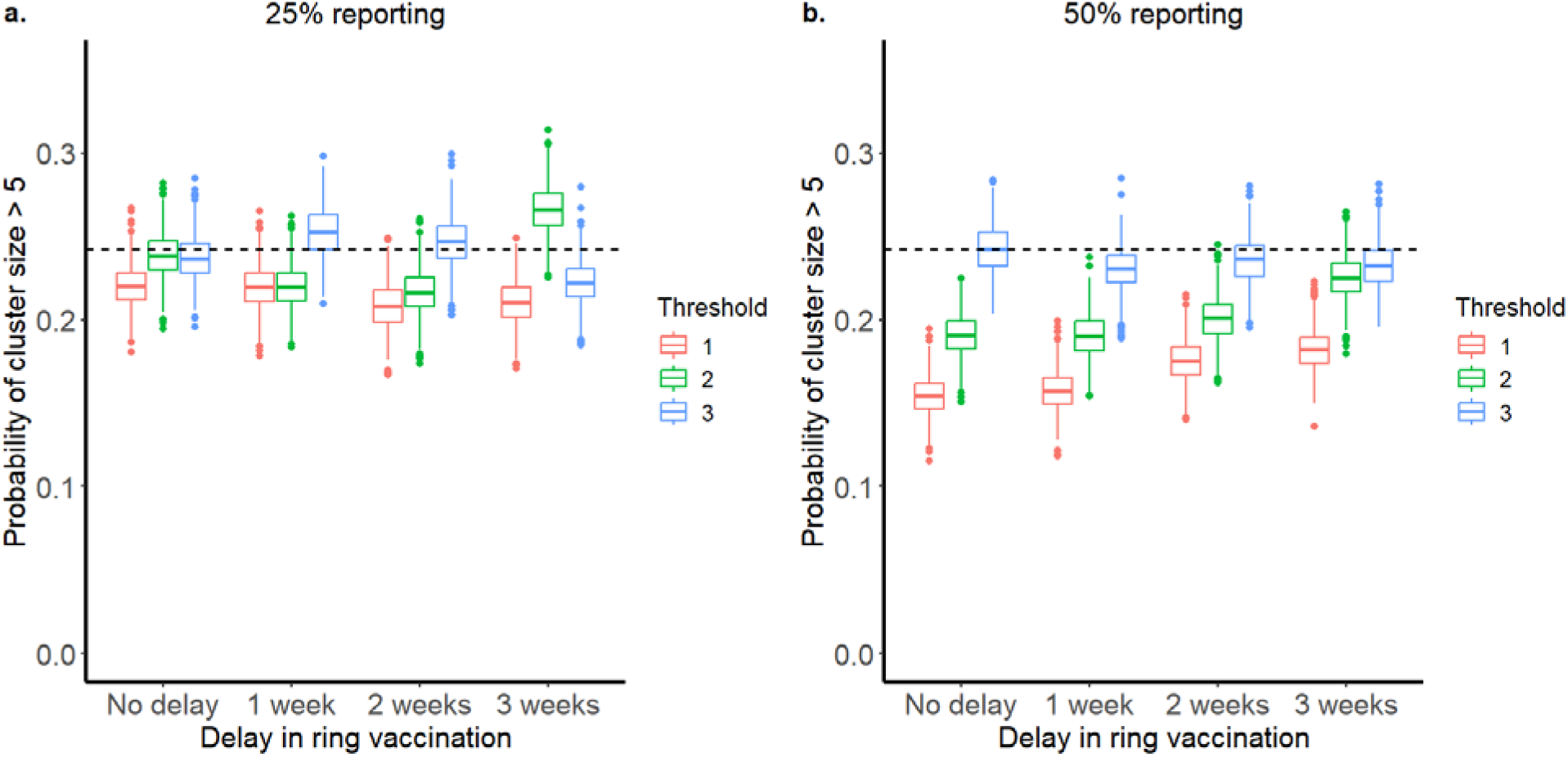
Probability of cluster size being greater than 5 when ring vaccination is triggered by different thresholds of reported infections. No sexual transmission is assumed, and the vaccination is done in households with reported infection. Fig. 3a. shows results for the household reporting of 25% and Fig. 3b. shows results for the household reporting of 50%. The dashed line represents the outcome from the baseline simulation with no vaccination.

### Vaccinating reported households and their non-household close contacts (between households) in the non-sexual transmission model

In this case, ring vaccination significantly reduces the probability of large clusters even with only 25% reporting (Fig. 4a). For example, with a reporting threshold of 1 and a 1-week delay in ring vaccination initiation, there is a 26% reduction in the risk of the outbreak size being greater than 5 when additional contacts of households are included for ring vaccination. The outcomes are improved with higher reporting (Fig. 4b). For example, when the reporting rate is increased to 50%, the relative risk reduction is 54% for a reporting threshold of 1 and a 1-week delay in ring vaccination initiation. Hence, the probability of occurrence of a large cluster can be significantly reduced if ring vaccination is performed for every reported infection within 1 week of reporting. Additionally, responding to every reported infection is better than responding after multiple infections are reported, even with a 2-week delay in ring vaccination initiation.

**Fig. 4.**
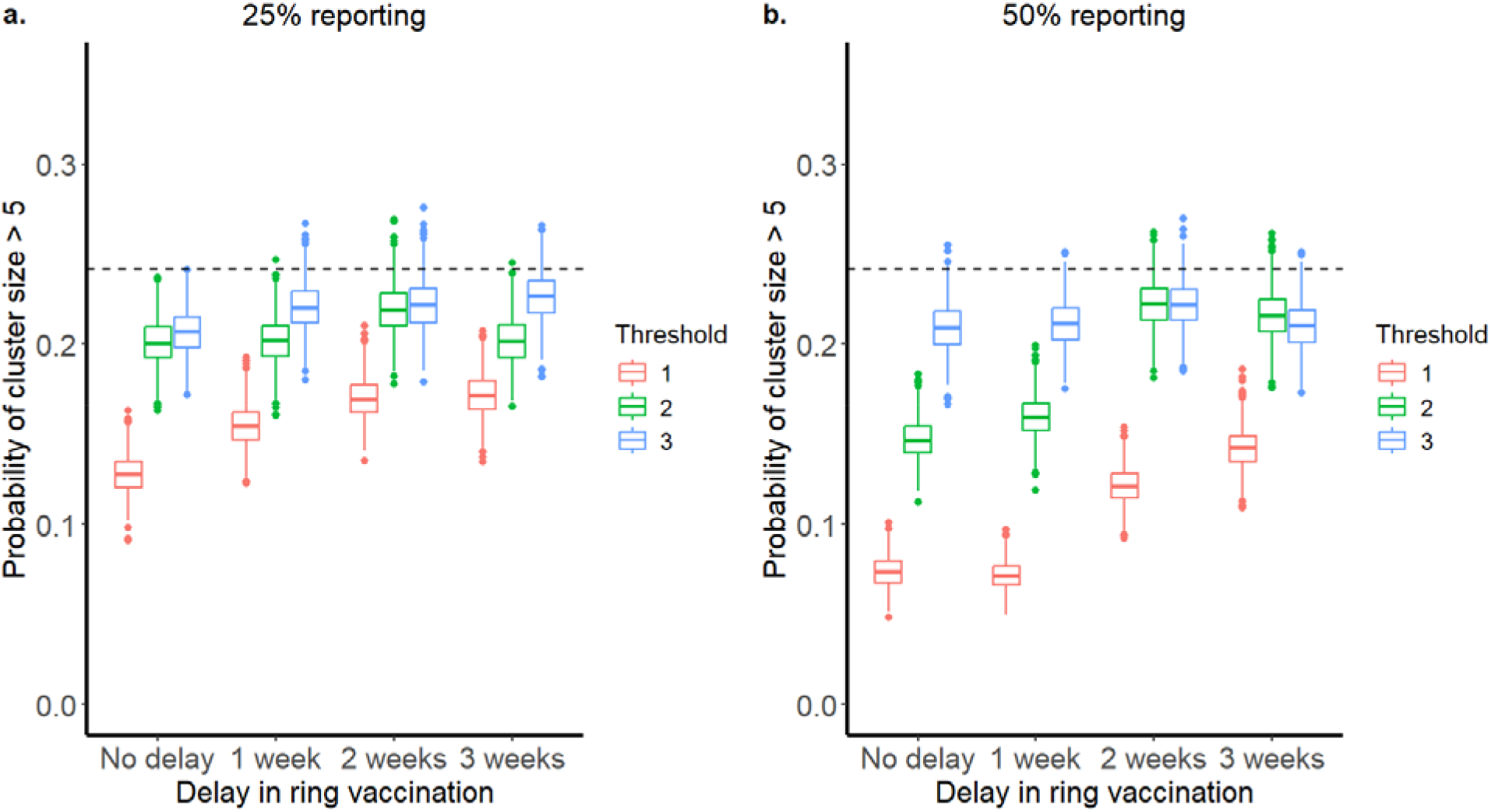
Probability of cluster size being greater than 5 when ring vaccination is triggered by different thresholds of reported infections. No sexual transmission is assumed, and the vaccination is done in households with reported infection as well as their non-household close contacts (between households). Fig. 4a. shows results for 25% of households reporting, and Fig. 4b. shows results for 50% of households reporting. The dashed line represents the outcome from the baseline simulation with no vaccination.

### Vaccinating only reported households in the non-sexual and sexual transmission model

When transmission via sexual network was added to the model, the baseline probability of cluster size being greater than 5 increased considerably (from 0.25 to 0.40, Fig. 3 and Fig. 5). Similar to Fig. 3, with 25% reporting, we do not see a significant reduction in the probability of the cluster size being greater than 5. Even with a 50% reporting, the probability decreases slightly only for the reporting threshold of 1, indicating that vaccinating only reported households may not be an effective strategy. On the contrary, comparison of mean outbreak size shows a significant reduction for the reporting threshold of 1, even with a delay of 3 weeks to initiate ring vaccination (Fig. S2).

**Fig. 5.**
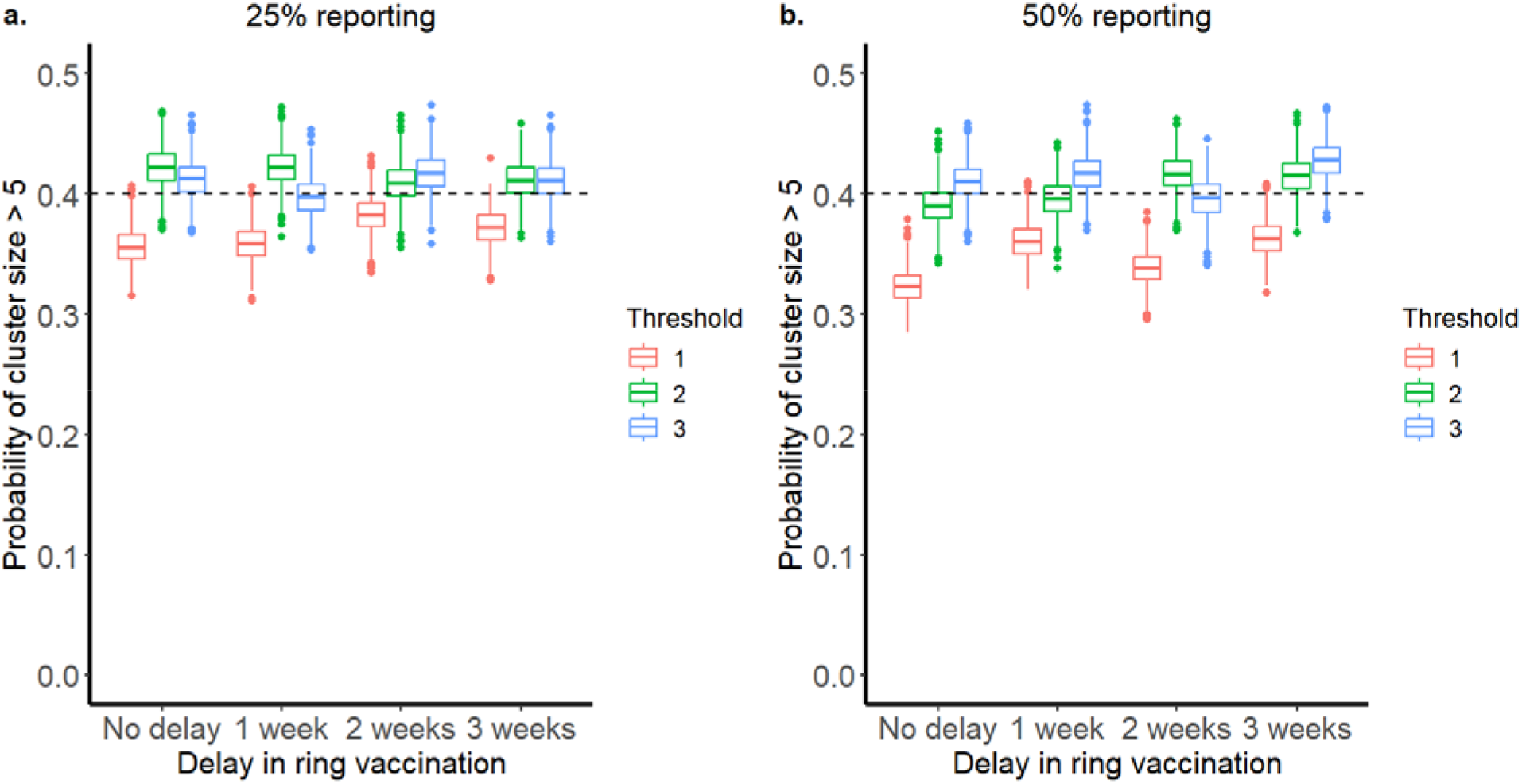
Probability of cluster size being greater than 5 when ring vaccination is triggered by different thresholds of reported infections. Sexual transmission, in addition to non-sexual close-contact transmission, is assumed, and the vaccination is done in households with reported infection. Fig. 5a. shows results for the household reporting of 25% and Fig. 5b. shows results for the household reporting of 50%. The dashed line represents the outcome from the baseline simulation with no vaccination.

### Vaccinating reported households and their non-household contacts in the non-sexual and sexual transmission model

When non-household close contacts were included for ring vaccination, the probability of observing a cluster size greater than 5 reduced significantly for immediate vaccination without any delayed initiation (Fig. 6a); this outcome was further improved with increased reporting rate (Fig. 6b). Response to every reported infection (i.e., reporting threshold=1) resulted in a lower probability of larger outbreaks, but this effect was diminished with a delay of 1 week or more with a reporting threshold of 2 or more cases. The comparison of relative risk reduction shows that improved reporting and responding to every reported infection provides the best result (Table 2). Additionally, we measured the mean outbreak size as a proportion of the total population that was infected. With a 50% reporting rate, the mean outbreak size was reduced by more than half for all response thresholds considered when ring vaccination was initiated without delay (Fig. S3). Delay to initiation increased outbreak size for all thresholds. The mean outbreak size for a 3-week delay in ring vaccination initiation for all reporting thresholds was smaller than the mean outbreak size for a reporting threshold of 2 and a 1-week delay, suggesting outbreak size is more sensitive to reporting threshold than delay in ring vaccination initiation. Under 50% reporting, the outbreak size was reduced by more than 20% for all delays considered when non-household close contacts were included in ring vaccination compared to the scenario when only within-household contacts were vaccinated (Table S1).

**Table 2.** Relative risk reduction with additional vaccination of contacts of infected households.

| Close-contact transmission | Delay in ring vaccination | 25% Reporting |  |  | 50% Reporting |  |  |
| --- | --- | --- | --- | --- | --- | --- | --- |
|  |  | Thresholds of reported infections for response initiation |  |  |  |  |  |
|  |  | 1 | 2 | 3 | 1 | 2 | 3 |
|  | No delay | 42% | 13% | 6% | 52% | 23% | 10% |
|  | 1 week | 26% | 6% | 4% | 54% | 16% | 0% |
|  | 2 weeks | 17% | 0% | 5% | 30% | 0% | 0% |
|  | 3 weeks | 13% | 15% | 0% | 21% | 0% | 0% |
| Close-contact and sexual contact transmission | No delay | 16% | 25% | 18% | 25% | 34% | 37% |
|  | 1 week | 5% | 8% | 0 | 30% | 6% | 6% |
|  | 2 weeks | 7% | 1% | 3% | 16% | 7% | 0 |
|  | 3 weeks | 2% | 0 | 0 | 6% | 4% | 4% |

**Fig. 6.**
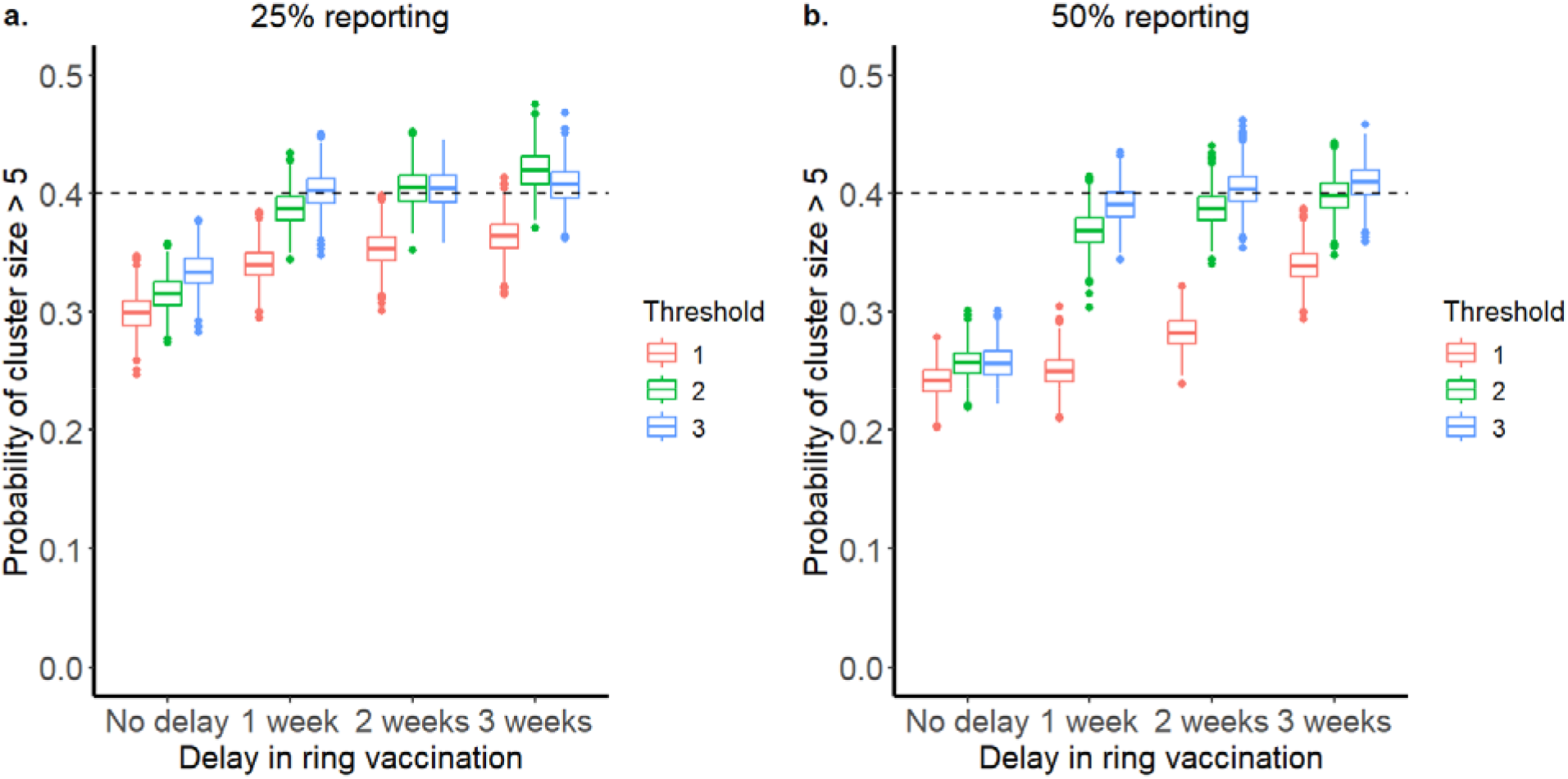
Probability of cluster size being greater than 5 when ring vaccination is triggered by different thresholds of reported infections. Sexual transmission, in addition to non-sexual close-contact transmission, is assumed, and the vaccination is done in households with reported infection and their close contacts. Fig. 6a. shows results for the household reporting of 25%, and Fig. 6b. shows results for the household reporting of 50%. The dashed line represents the outcome from the baseline simulation with no vaccination.

## Discussion

Before the global outbreak of 2022 due to clade IIb, MPXV transmission did not exhibit extended human-to-human transmission. Rather, it was limited to close contacts in space and time. The longest reported transmission chain for clade I mpox was seven generations long [14]. During 2023-2025, the DRC has reported multiple provincial-level outbreaks of clade I, partially driven by sexual transmission [4]. Understanding what resources are needed and how we can control local transmission during different stages of outbreaks is needed to stop transmission from spreading to additional regions. Recently, Savinkina *et al*. explored the benefits of mass vaccination against mpox in controlling the outbreak in DRC [29]. In this study, we used modeling to investigate the effectiveness of ring vaccination with the constraints of limited resource availability and limited surveillance. Even with these limitations, we show that ring vaccination can be an effective tool in managing outbreaks at the local community level.

Low- and middle-income countries often have limited resources to deal with public health threats. Implementation of ring vaccination for each spillover or importation case in an area could be an effective strategy to control outbreaks early and reduce their impact on the community. When the disease is rare, it requires far fewer vaccines and resources to implement ring vaccination compared to mass community vaccination. Mpox is still a relatively rare disease. For example, even in the global outbreak of 2022, only a small proportion of the entire population was at risk of being infected [30]. Additionally, even in endemic areas, the disease disproportionately affected certain populations. For example, data from the current outbreak in DRC suggest that clade Ia predominantly affects children under 15 years old, whereas clade Ib has so far predominantly transmitted via sexual contact in adults [31]; however, change may occur as the disease spreads [32]. Therefore, targeted ring vaccination can be effective against mpox. However, in disease hotspot areas where the spillovers are frequent, this may be difficult to implement because of the resources required to deal with each spillover event. Initiating a response after multiple infections can potentially reduce the frequency of the responses since some of the spillovers will not cause onward transmission and hence would not trigger ring vaccination initiation. At the same time, if surveillance is weak and the reporting rate is low, many cases can be missed that can potentially lead to larger outbreaks. For example, when mpox is driven mainly by non-sexual close-contact transmission, we have shown that waiting for three or more infections in a household to be reported is ineffective, as it does not reduce the probability of a large cluster. However, if ring vaccination is performed within two weeks of reporting two cases, then the probability of observing a large cluster reduces significantly. While performing ring vaccination for each report of spillover results in the best-case scenario, acting after two reported infections can still be helpful while also reducing the frequency of the responses occurring.

The success of ring vaccination is tied to strong surveillance for case identification, contact tracing of exposed individuals, and fast mobilization of public health resources. The reporting rate for mpox in many rural areas of DRC has traditionally been low [10], and active surveillance has been limited to a few locations. Similarly, many of the cases occur in remote areas with limited accessibility. Hence, responding to reports can be delayed significantly. Our model shows that for non-sexual close contact transmission, while a delay in response reduces the effectiveness of ring vaccination, aggressive contact tracing can still reduce the likelihood of a large cluster, even with limited reporting. In most scenarios, responding to every report within two weeks significantly reduces the likelihood of large outbreaks. This supports the current recommendation of the U.S. Centers for Disease Control and Prevention for post-exposure vaccination within 4 days in ideal conditions and up to within 14 days [26]. We further show low and high ranges of ring vaccination effectiveness depending on the identifiability of non-household contacts. The effectiveness of ring vaccination in real-world scenarios may lie within this range, as a certain proportion of non-household contacts may not be identified or cannot be accessed. For example, close to 20% of more close contacts were not identified or refused vaccine even when identified during the 2019 Ebola outbreak in DRC [33].

Following continuous spillovers and stuttering transmissions, new strains can arise that have higher transmission rates and/or modified epidemiology. The Clade I outbreak of 2023 and 2024 shows a different epidemiology compared to previously reported outbreaks in the DRC. Previously, most of the human-to-human transmission was due to close contact within households. It is still believed to be a primary transmission route for Clade Ia that is affecting the western part of the DRC, although sexual transmission has also been reported in some cases [32]. Clade Ia has also been confirmed in neighboring countries, including the Republic of Congo and the Central African Republic [30]. However, a small cluster involving transmission through sexual contact in men who have sex with men (MSM) population was observed in early 2023 [7]. Further, most suspected cases in Kamituga, a mining town in eastern DRC, were linked to commercial sex activities involving men and women [8,34]. The strain of MPXV causing the outbreak in Kamituga - clade Ib and clade Ia in Kinshasa have APOBEC3 mutations similar to the clade IIb strain that caused the global outbreak of 2022, which are indicative of persistent human-to-human transmission. Recent evidence suggests co-circulation of Clade Ia and Clade Ib MPXV in Kinshasa [35] which may also be true in other provinces, resulting in complex epidemiological trends. As of early September 2024, cases due to clade Ib MPXV have been confirmed in neighboring countries, including Burundi, Rwanda, and Uganda, possibly imported due to the mobile nature of commercial sex workers and clients in the region [30]. Travel-associated cases are also confirmed in several countries, including Thailand, Sweden, India, Germany, UK, USA, and Australia, highlighting its potential for global impact. While commercial sex workers could be targeted for vaccination if issues related to stigma and sex work legality can be overcome, ring vaccination in sexual networks is still difficult to implement since sexual contacts are hard to trace given the frequency of anonymity. Hence, we assessed the effectiveness of ring vaccination of only non-sexual contacts, but in the presence of additional sexual transmission. We found that ring vaccination of only within-household contacts, failing to vaccinate non-household non-sexual contacts, does not reduce the likelihood of a large cluster. However, it can still reduce the overall outbreak size if ring vaccination is done early and for every reported infection. When including non-household nonsexual contacts for ring vaccination, the likelihood of a larger cluster is reduced with a much smaller overall outbreak size. Hence, although ring vaccination may not be effective in controlling infections at the source, responding to every reported infection can still reduce the overall impact on the community.

The model has several limitations, often made for computational efficiency. We fitted the model with data from 2013 to 2017. However, some of the parameters may have changed, including the transmission rate, which has led to a significant increase in case numbers in 2023-2024, which is mainly due to several environmental factors – e.g., the sexual network in which the virus was introduced -, and not related to genetic changes in the virus. Additionally, we kept the individuals per household constant, and we do not consider age structure in the population. Heterogeneities in household size and composition may also play a role in determining the outbreak size, especially for scenarios driven by non-sexual close contact transmission [36,37]. We further assume that individuals who are vaccinated acquire immunity immediately, which may not be true. Additionally, post-exposure vaccine efficacy may be different from previous estimates of vaccine efficacy. New studies of post-exposure vaccine effectiveness in DRC are currently proposed but have not yet been completed. The number of sexual partners per day and the probability of transmission given sexual contact are not well known in many DRC settings, and these uncertainties can change the results presented here. Additionally, we only examined the effect of ring vaccination during the early stages of potential outbreaks. The recommendations and results shown here may not apply to cases when the disease is already widespread in a community. However, this model framework is suitable for informing decision-making at a local community scale. Other models are currently available that can help in decision-making at a broader regional scale [38].

Zoonoses often follow multiple stages where spillover can lead to either no further cases, stuttering transmission, or sustained human-to-human transmission [39,40]. Each stage presents unique opportunities and challenges to address disease spillover and transmission risk. Identifying early stages of spillover requires a strong surveillance system, whereas preventative measures such as vaccination provide protection from pathogens with stuttering and sustained transmission potential. Preventing spillovers or imported cases with different transmission potential is crucial during the early phases of the outbreak. For many zoonoses that have vaccines available, proactive measures of community vaccination can provide good protection against disease spillover and transmission. However, for neglected tropical diseases such as mpox, vaccine availability in remote areas is a serious issue due to logistic difficulties for distribution. Although reactive measures, such as ring vaccination with far fewer vaccines, can be effective in controlling the transmission, other challenges, such as contact tracing, accessibility, and case identification, can influence the effectiveness of ring vaccination. In this study, we presented scenarios where ring vaccination can be effective in reducing the likelihood of a larger outbreak. These analyses can inform public health officials on whether and how to respond to spillover or imported cases in a local community. For positive outcomes of reduced outbreak size and likelihood of large clusters, we show a trade-off between the delay in ring vaccination initiation and the reporting threshold required to trigger ring vaccination. Hence, depending on the local scenario, one of these two aspects can be prioritized. When a delay in ring vaccination initiation is unavoidable, responding to every reported infection gives better outcomes. On the other hand, if ring vaccination can occur rapidly in a community, waiting for multiple reported infections might be preferable. Hence, our study provides practical approaches for managing outbreaks in resource-limited communities.

## Data Availability

All data produced in the present study are available upon reasonable request to the authors.

## Acknowledgements

The findings and conclusions in this report are those of the authors and do not necessarily represent the official position of the Centers for Disease Control and Prevention.

## Author Contributions

AP, IS, and YN contributed to the study conception and design. Analysis was performed by AP. The first draft of the manuscript was written by AP, and all authors commented on the previous versions of the manuscript. All authors read and approved the final manuscript.

