## Supplemental Figures for "Optimizing vaccination strategies for mpox control in endemic areas: Modeling insights from the Democratic Republic of Congo"

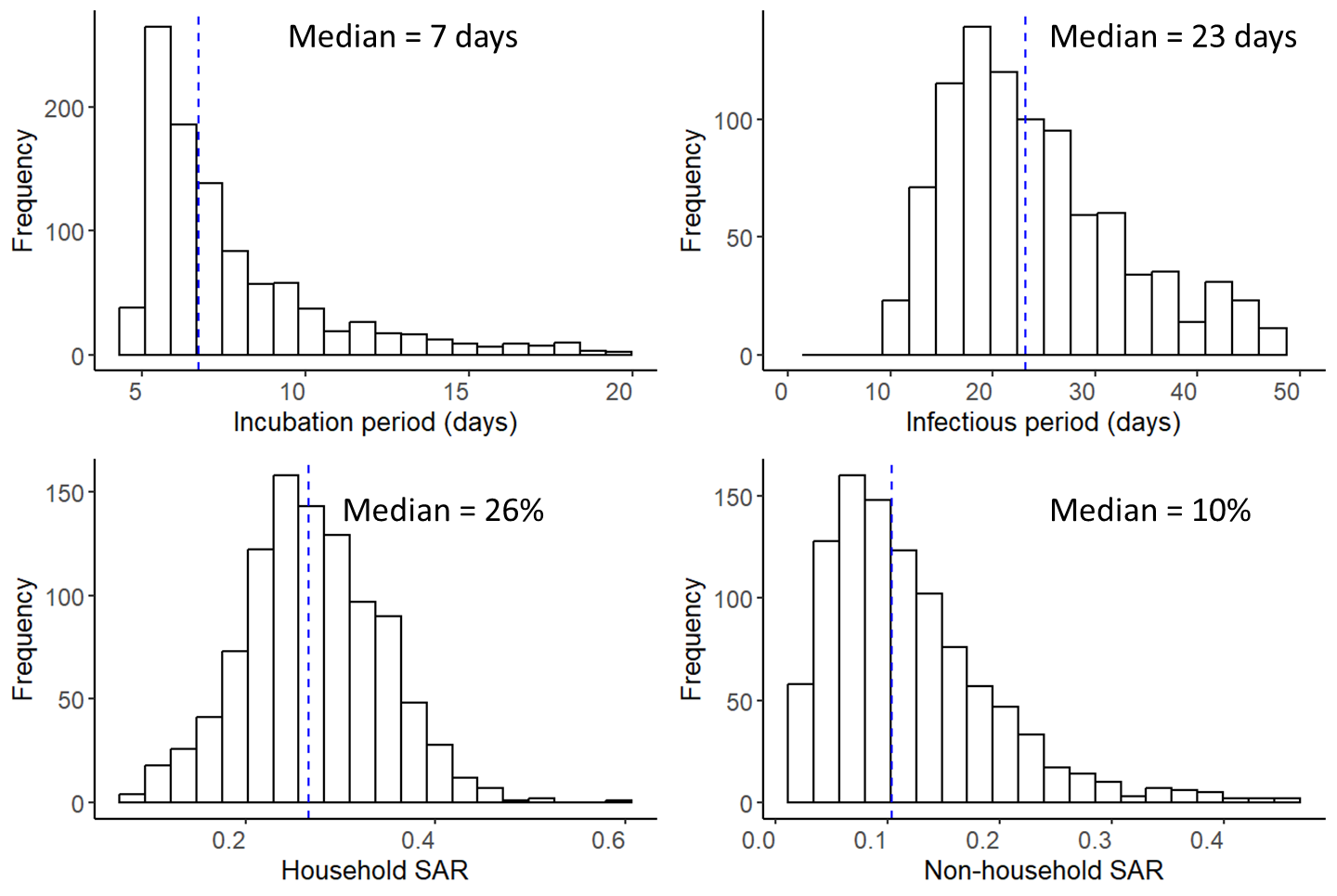

Fig. S1. Estimated parameter distribution and median values.

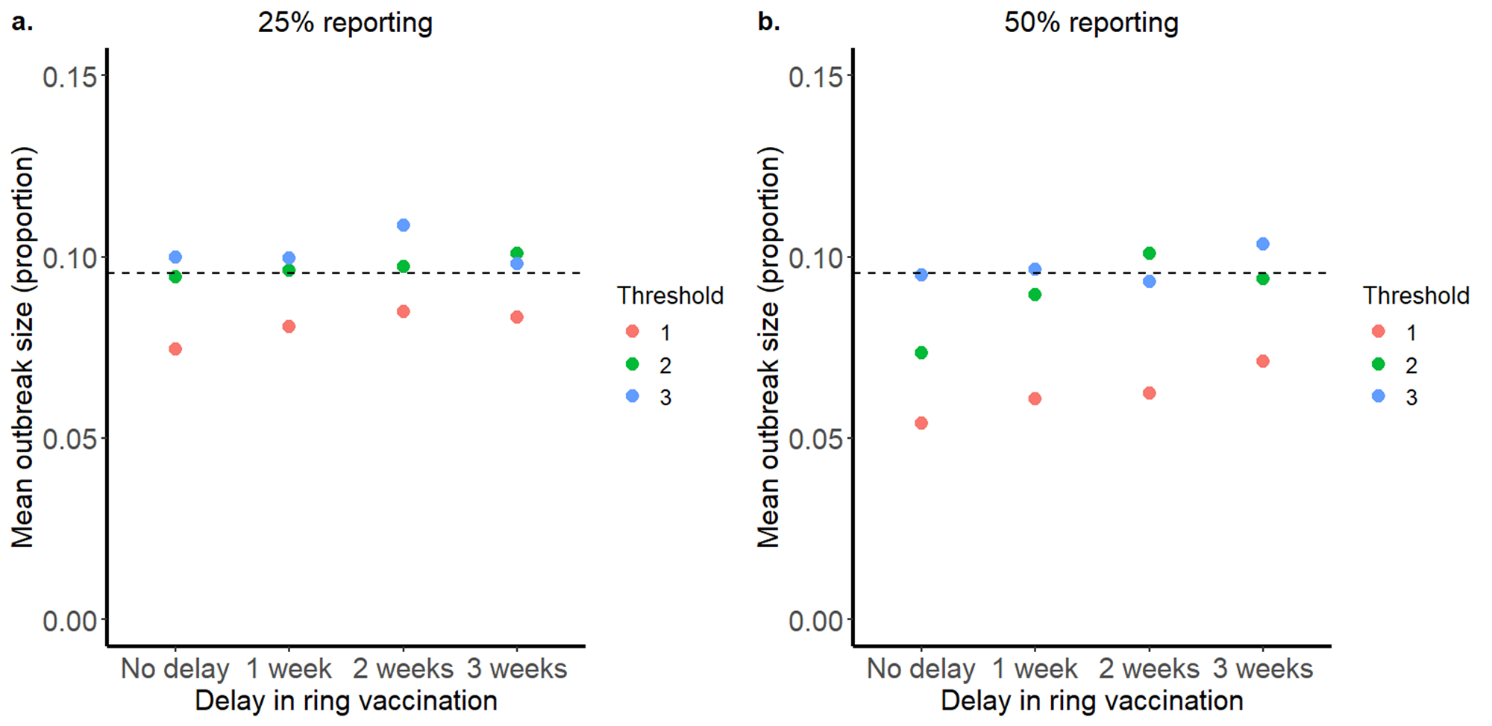

Fig. S2. Mean outbreak size as a proportion of the total population infected for household vaccination from 1000 simulation outputs. Sexual transmission was included in this scenario. Only the reported households are vaccinated. The dashed line shows the baseline outbreak size.

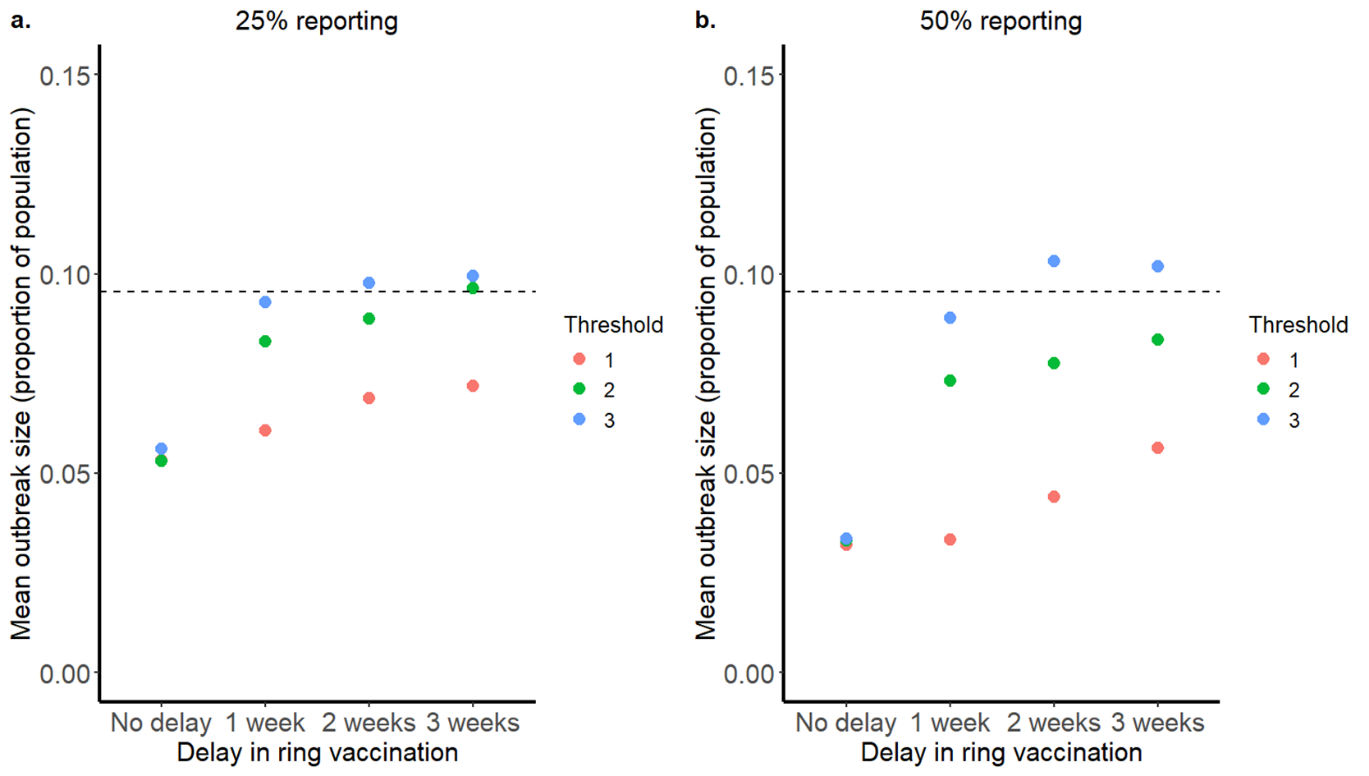

Fig. S3. Mean outbreak size as a proportion of the total population infected for household vaccination from 1000 simulation outputs. Sexual transmission was included in this scenario. The reported households and their close contact households are vaccinated. The dashed line shows the baseline outbreak size.

|  | **25% Reporting** | | | **50% Reporting** | | |
| --- | --- | --- | --- | --- | --- | --- |
|  | **Thresholds of reported infections for response initiation** | | | | | |
|  | 1 | 2 | 3 | 1 | 2 | 3 |
| No delay | 28% | 43% | 43% | 41% | 55% | 64% |
| 1 Week | 25% | 14% | 7% | 45% | 18% | 7% |
| 2 Weeks | 19% | 9% | 10% | 29% | 23% | 0 |
| 3 Weeks | 14% | 4% | 0 | 21% | 11% | 0 |

Table S1. Relative reduction in mean outbreak size with additional vaccination of contacts of reported households compared to vaccination of reported households only.
